# cfDNA UniFlow: A unified preprocessing pipeline for cell-free DNA data from liquid biopsies

**DOI:** 10.1101/2024.06.01.24308210

**Authors:** Sebastian Röner, Lea Burkard, Michael R. Speicher, Martin Kircher

## Abstract

**Background:** Cell-free DNA (cfDNA), a broadly applicable biomarker commonly sourced from urine or blood, is extensively used for research and diagnostic applications. In various settings, genetic and epigenetic information is derived from cfDNA. However, a unified framework for its processing is lacking, limiting the universal application of innovative analysis strategies and the joining of data sets.

**Findings:** Here, we describe cfDNA UniFlow, a unified, standardized, and ready-to-use workflow for processing cfDNA samples. The workflow is written in Snakemake and can be scaled from stand -alone computers to cluster environments. It includes methods for processing raw genome sequencing data as well as specialized approaches for correcting sequencing errors, filtering, and quality control. Sophisticated methods for detecting copy number alterations and estimating and correcting GC-related biases are readily incorporated. Furthermore, it includes methods for extracting, normalizing and visualizing coverage signals around user defined regions in case-control settings. Ultimately, all results and metrics are aggregated in a unified report, enabling easy access to a wide variety of information for further research and downstream analysis.

**Conclusions:** We provide an automated pipeline for processing cell-free DNA sampled from liquid biopsies, including a wide variety of additional functionalities like bias correction and signal extraction. With our focus on scalability and extensibility, we provide a foundation for future cfDNA research and faster clinical applications.

**Issue Section:** Technical Note

**Availability and implementation:** Source code and extensive documentation is available on our GitHub repository (https://github.com/kircherlab/cfDNA-UniFlow).

## Introduction/Background

Cell-free DNA (cfDNA) is found in many bodily fluids like blood plasma and urine [1]. It is believed to be primarily derived from natural degradation processes during cell turnover [2]. However, the proportion of cell-types and tissues contributing to cfDNA changes in the context of certain physiological conditions or disease processes [3,4]. Thus, signals in cfDNA might serve as relevant biomarkers in health and disease. Collecting cfDNA in so-called liquid biopsies (Fig. 1) is considered non-invasive and led to an increased research interest in the biomedical field for using cfDNA in allograft (i.e., donor organ) rejection, prenatal testing and diagnostics, as well as disease detection and health monitoring [5] (especially for cancer).

**Figure 1.**
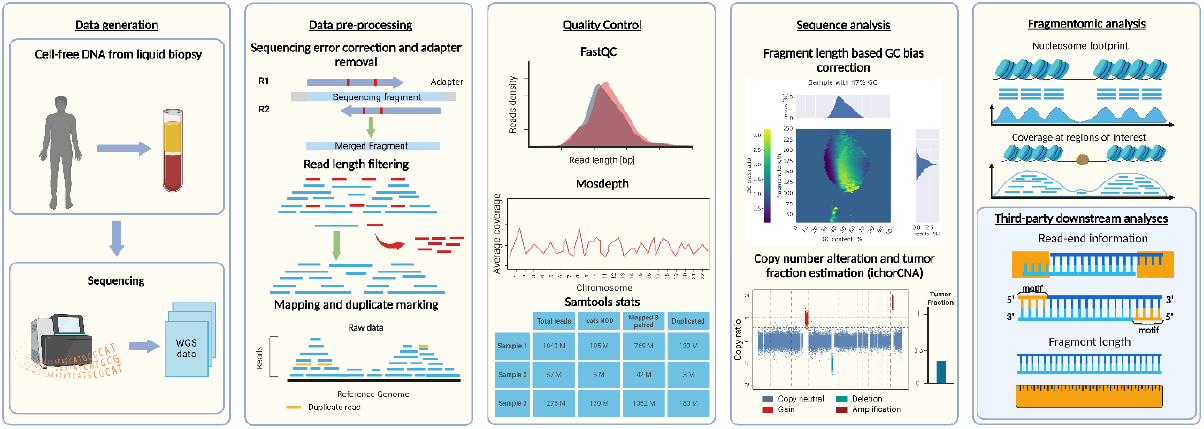
Overview of cfDNA analysis. The leftmost panel depicts data generation by liquid biopsy sampling followed by library preparation and sequencing. The second panel shows the entry point of cfDNA Uniflow. It displays the core functionality of merging reads/removing adapters, length filtering, mapping to a reference genome and duplicate marking. Sample quality control is shown in the third panel and for example performed using FastQC, Mosdepth and SAMtools stats. The fourth panel shows optional steps of GC bias correction and estimation of copy number alterations and tumor proportion. Finally, results are aggregated, for example in a report and used for downstream analyses (fifth panel). Figure created with Biorender.

Over the last years, many approaches have been developed to extract information from cfDNA samples for various applications. Methods range from identifying allelic differences at known disease markers, detection and tracking of mutations[6] and copy number alterations (CNAs) in tumor cells [6], and DNA fragmentation differences [3,8,9] to measuring methylation state [10–12]. While these methods exploit different signals, all rely on the precise quantification of read distributions, and slight changes in read recovery affect their results (Fig. 1).

Therefore, consistent data quality is the primary requirement for developing these new diagnostic methods (Fig. 1). Even though sample handling is constantly streamlined, individual differences of sample donors, and logistic factors like time of sample collection, duration, conditions of storage, and further preanalytical handling are challenging to fully control in a clinical context, but have been shown to affect the quality of cfDNA samples [13–16]. Additionally, detecting signals of interest (e.g., from circulating tumor DNA, ctDNA) in a background mainly derived from hematopoietic cells [16] is not trivial, emphasizing the need for optimal data quality.

One way to mitigate some preanalytical effects and technical biases introduced during sequencing of cfDNA samples is to include specialized correction and sampling steps during computational processing of the data (Fig. 1). Even though the need has been identified previously in the field of cfDNA, community standards are still lacking for preprocessing genome sequencing data from cfDNA [7,8,18– 23].

One reason for the lack of dedicated cfDNA pipelines, might be that many publications in the field are focused on the downstream analysis like the classification of disease samples, relying on unpublished in-house pipelines for data processing. Further, important correction steps are often tailored towards specific features and tightly integrated in downstream analysis pipelines, making it difficult to generalize and transfer them to new projects [7,8]. Nevertheless, there have been some approaches trying to address the need for community standards. A notable one is the FinaleDB project, which aggregates cfDNA samples from multiple sources, processes them in a uniform manner and provides fragment coordinates via a web portal [23]. To protect the privacy of patients, the data is anonymized during processing, removing all sequence information and making it unsuitable for analyses not focused on fragmentation patterns. Additionally, this pipeline does not address issues of batch and bias correction, which might be most relevant in such aggregation efforts. The project getting closest to setting a community standard for processing samples not just for fragmentomics applications, is called cfDNApipe. It combines many useful tools for basic processing of normal and bisulfite converted DNA sequences. The utility functions range from generation of summary statistics, GC-bias correction tailored towards CNV detection and extraction of a limited number of features [25]. However, the software seems to be designed for single computer use, lacking many of the features provided by a full-fledged workflow management system, making it hard to scale analysis in different environments, like compute clusters. Moreover, the design does not allow for easy integration of new functionalities, creating the need for either an additional workflow management system or extensive modification of the original code.

Technical biases and missing community standards cause several drawbacks for the field. First, users rely on standard processing pipelines from other fields, which might not be suitable for specific analyses. They might also feel the need to develop their own pipelines by selecting appropriate tools and tuning parameters optimized on the available set of samples. Second, it adds additional overhead when comparing across multiple studies. Here, researchers are frequently required to work with the original processing of each site, potentially introducing technical biases in the analysis. Alternatively, reprocessing data from multiple sites can reduce technical biases between studies but creates an additional computational and organizational burden (incl. access to raw and protected genetic data). Third, it can be hard to keep track of all sample-level information when building analysis pipelines using many samples, mainly when information gets scattered across many samples and files.

To jointly address several of these problems, we developed an easy-to-use unified preprocessing workflow for cell-free DNA written in Snakemake. It combines a curated list of tools for processing genomic cfDNA samples, custom tools for reducing technical biases, and tools for estimating additional characteristics like copy number states. Our pipeline is implemented with high configurability, scalability from single computers to high-performance compute clusters, and a sophisticated reporting system.

## Overview and implementation

### Implementation

We implemented the cfDNA UniFlow workflow in the popular workflow management system Snakemake [25]. This makes it easy to scale the workflow in different computing environments and allows for parallel processing of multiple samples. Further, most of the rules are implemented to enable multiprocessing and efficiently utilize multiple cores for each task. Conveniently, default resources like genome references or standard adapter files can be downloaded, if not configured to point to already available resources. A detailed overview of the workflow is available in Figure S1. Briefly, cfDNA UniFlow covers three parts between data generation and downstream analysis: data pre-processing, quality control and utility functions (Figure 1).

### Preprocessing

The core preprocessing steps (Fig 2., components depicted in blue) expect FASTQ files as input. Alternatively, existing alignments (BAM files) can be provided for reprocessing. In the latter case, the workflow automatically converts these to FASTQ files using SAMtools [26]. Afterwards, reads can be merged with NGmerge [27], which also removes sequencing library adapters and corrects sequencing errors and ambiguous bases based on the read-overlap consensus. Reads that were not merged, can be postprocessed using NGmerge adapter removal mode and can be included in the mapping process. Alternatively, the merging step can be skipped and reads will only be trimmed using Trimmomatic. Prior to mapping with bwa-mem2 [29], reads are further filtered based on their length, excluding reads that are shorter than a configurable threshold. Finally, duplicate reads are marked (SAMtools markdup) before the BAM files of the samples are passed to the next step.

**Figure 2.**
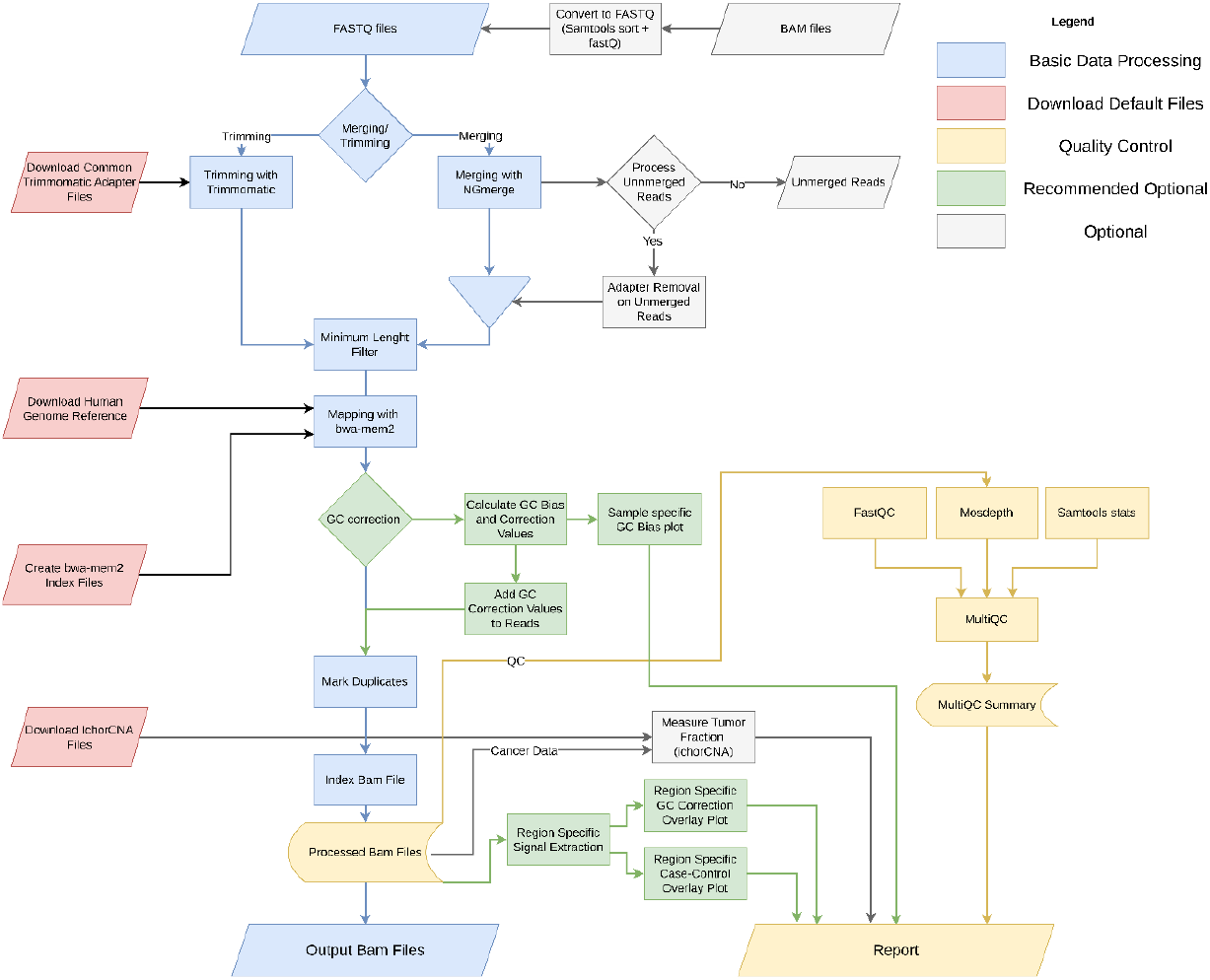
Overview of unified cfDNA preprocessing workflow. Functionalities are color coded by task. Blue boxes contain the core functionality of cfDNA Uniflow. Red boxes represent rules for the automatic download of public resources. Yellow boxes summarize the Quality Control and reporting steps. Finally, grey and green boxes are optional steps, with green boxes being highly recommended.

### Quality Control

In the Quality control (QC) step (Fig. 2, components depicted in yellow), general post-alignment statistics and graphs are calculated for each sample with SAMtools stats [26] and FastQC [30]. Additional information on sample-wide median coverage and coverage at different genomic regions is calculated via Mosdepth [31]. The QC results are aggregated in an HTML report via MultiQC [32] and an example is shown Figure S2.

### Signal Extraction

In the last step, additional utility modules (Fig. 2, components depicted in green) can be configured and executed. This includes our in-house GC bias estimation and correction methods (https://github.com/kircherlab/cfDNA_GCcorrection), an extension of the method described by Benjamini & Speed [17]. As fragmentation in cfDNA is driven by natural degradation processes, libraries constructed from liquid biopsies tend to have fragments of a wide range of lengths and do not follow the original assumption that length is well-approximated by the mean fragment length. Therefore, we estimate the expected fragment distribution by sampling regions along the reference genome, counting all possible fragments for a specified range of fragment lengths and sorting them in bins of their GC content. Afterwards, we measure the sample specific fragment distribution in the same regions, scale them and compare them to the theoretical distribution. Based on the ratio of observed and expected, we calculated correction values for each fragment length and GC content. The resulting weights are attached to the reads as tags, which can be used for a wide variety of downstream signal extraction methods, while preserving the original read coverage and fragmentation patterns. We provide specialized signal extraction routines to extract coverage derived signals using read weights. Further, we included the widely used tool ichorCNA [6], to identify copy number alterations and estimate tumor fraction. An example of the output is available in Figure S3.

### Reporting

Finally, all information provided by the previous steps is aggregated in a comprehensive HTML report. This includes summary statistics on workflow execution provided by Snakemake, and plots and summary statistics produced in the quality control steps. Additional information from optional steps includes a general estimation of sample-specific GC bias parameters (Figure S4), the effects of GC bias correction in user defined regions (Figure S5) and plots on copy number alterations created by ichorCNA. Finally, case-control plots are generated and included, if more than one class of samples is provided (Figure S6).

## Results

To test and showcase cfDNA Uniflow, we use three exemplary cfDNA samples ( healthy H01, breast cancer B01, prostate cancer P01) with different conditions and average GC contents from the European Genome-Phenome Archive Study EGAS00001006963. Each sample was converted to FASTQ files and processed in our pipeline with standard parameters for human reference build GRCh38/hg38. As userdefined regions of interest, we selected 10,000 binding sites of LYL1, a transcription factor (TF) associated with hematopoietic cells [33], and GRHL2, an important pioneer TF for epithelial cells [34– 36] playing a role in a wide variety of cancer types [37–41]. Both TFs are especially suited due to their association with expected tissue contributions in our samples and because they have high GC content binding sites.

This can be seen in Figure 3, which shows coverage overlays centered on LYL1 binding sites and illustrates the global and regional effects of GC biases in the respective samples. The healthy sample H01, with an average GC content of 45%, shows a balanced global GC profile (Fig. 3a) and, accordingly, the GC bias correction shows almost no effects on the composite signal. We see the strongest drop of coverage at the TF binding site, expected for a sample of mainly hematopoietic origin where many LYL1 binding sites are expected to be accessible to the TF. B01, a breast cancer sample with an average GC content of 38%, shows an overrepresentation of fragments with GC content lower than the genome average and an underrepresentation of fragments with higher GC content (Fig. 3b). This leads to a distortion of the composite coverage signal around the LYL1 binding sites. Without GC correction, the drop in coverage would be overestimated. After correction, coverage at the site is closer to the coverage of the surrounding regions, consistent with an expected signal dilution compared to the healthy sample (Fig. 3a) due to a higher contribution of non-hematopoietic cell-types in this cancer sample (Fig. 3b). The same should be true for sample P01 (Fig. 3c), a prostate cancer sample with an average GC content of 45%. However, the global GC bias profile (right panels) show the inverse trend to sample B01, with a shift of fragment distribution towards a higher GC content. Unsurprisingly, the signal around the binding sites is distorted towards higher coverage prior to the GC correction (i.e., suggesting that the TF binding sites are not accessible). After GC correction, the signal looks similar to the one shown for B01, less open than the healthy sample and consistent with an increased contribution of non-hematopoietic cell-types. Global effects of GC bias correction on fragment distribution and a comparison to two other fragment-based are provided in the Supplement (Figure S7).

**Figure 3.**
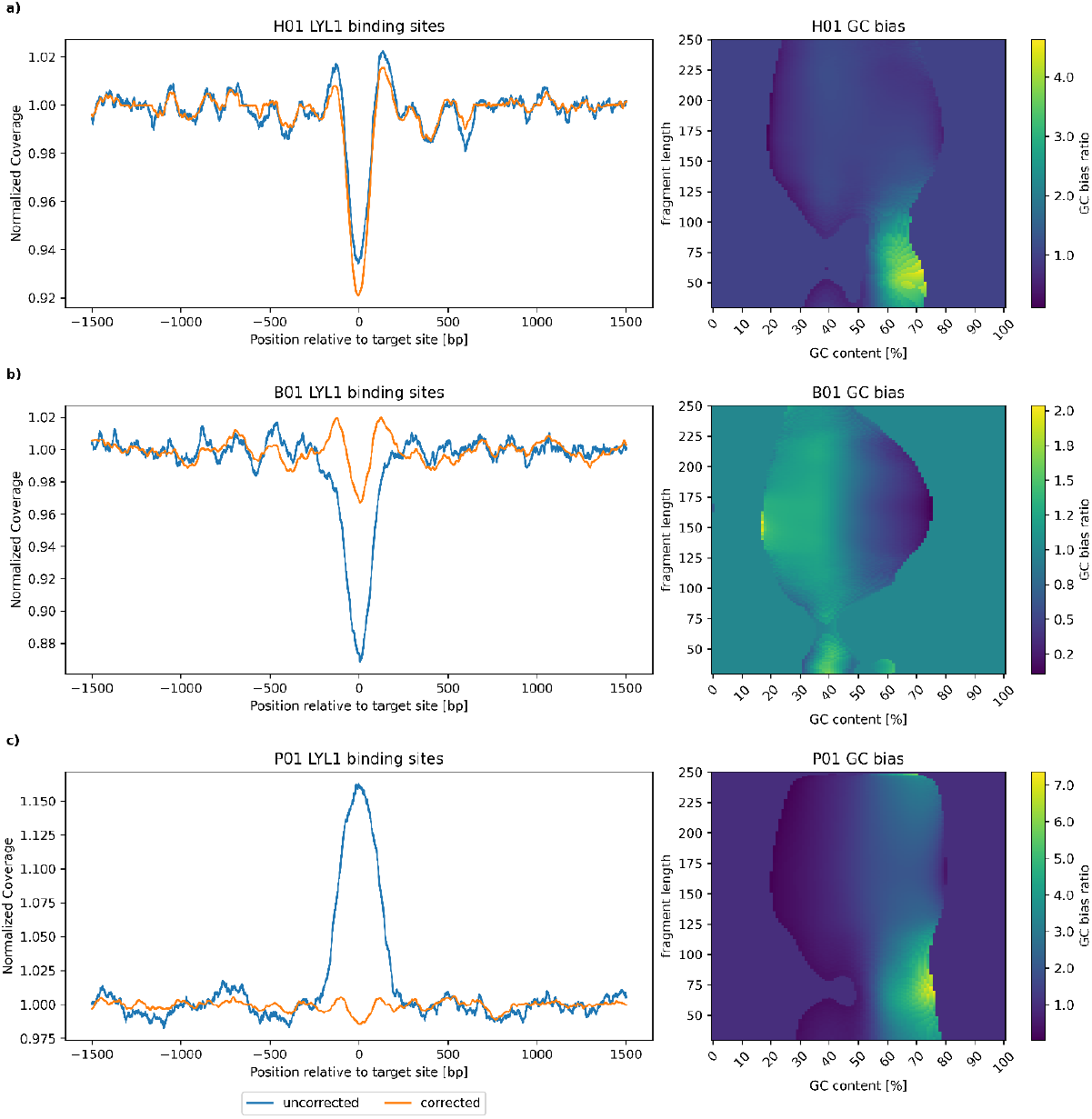
Effects of GC bias on regional and global scale. Composite coverage signals of 10,000 LYL1 transcription factor binding sites (left) and global bias profiles (right) for three cfDNA samples are shown. a) Signals and profile for a healthy sample (H01) with an average GC content of 41%. The GC profile (right) is relatively balanced between observed and expected fragments. Respectively, the GC bias corrections have only minor effects on the composite coverage signal (left). b) GC bias effects for a breast cancer sample with an average GC content of 38%. The global GC profile shows an overrepresentation of lower GC content fragments (brighter color) and an underrepresentation of higher GC content fragments (darker color). This results in an underestimation of coverage (overestimation of accessibility) at the LYL1 binding sites. After GC correction the signal is closer to the surrounding coverage, consistent with lower relative contributions of hematopoietic cells and fewer open sites. In contrast, c) shows the GC bias effects of a prostate cancer cell with an average GC content of 45%. The global GC profile is skewed towards a higher GC content, leading to an overestimation of coverage around the LYL1 binding sites. After GC correction, the signal is closer to the surrounding coverage, indicating lower contributions of hematopoietic cells with accessible LYL1 sites.

In addition to the GC bias plots for individual samples, we provide case-control plots for comparing sample classes with a control in the same plot. In our example, the healthy sample H01 would be the control and we are comparing samples on the LYL1 and GRHL2 sites. As noted, the expected signal around LYL1 binding sites is a drop in coverage for samples mainly derived from hematopoietic cells. When the contribution of non-hematopoietic cell-types, in which LYL1 is not expressed, increases, we expect to see a relative increase in coverage around the binding sites. Accordingly, the signals shown for our three test samples (Fig. 4a) are in line with that expectation. For GRHL2, we expect the opposite signal. As healthy samples should not include many contributions from tissues with high GRHL2 activity, the expected coverage signal should be similar to the surrounding regions. In contrast, samples with high contributions of cancer-derived DNA should show a drop in coverage, indicative of higher accessibility of the TF binding sites (Fig. 4b).

**Figure 4.**
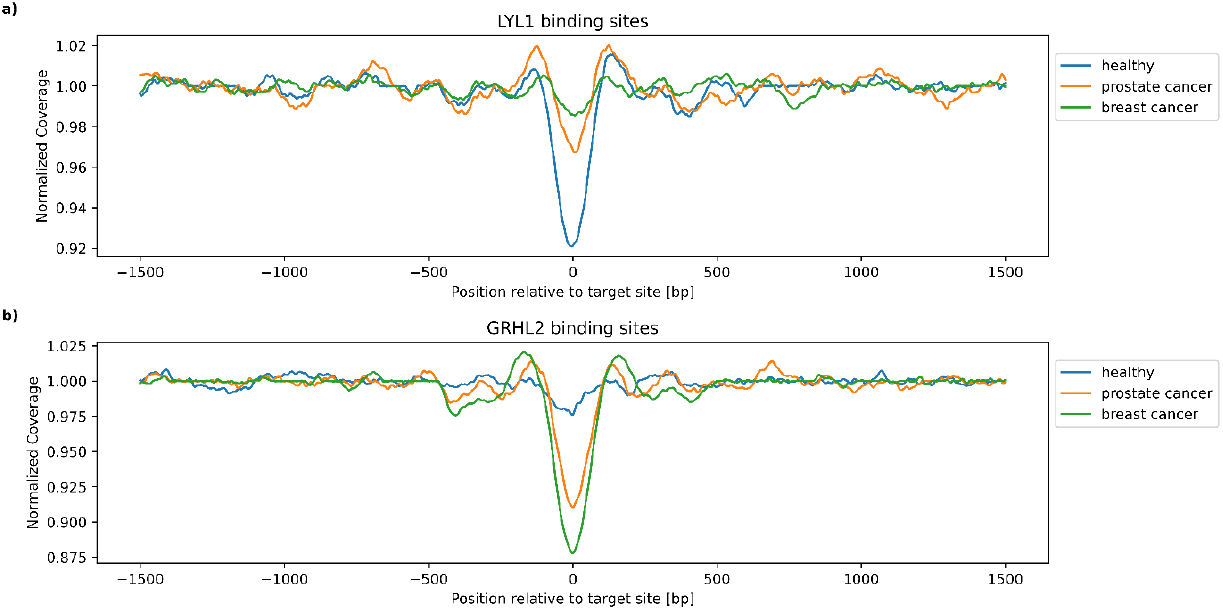
Case-control plots around GRHL2 and LYL1 binding sites. a) GC corrected composite coverage signals around 10,000 centered LYL1 transcription factor binding sites. The healthy sample (H01) shows lower relative coverage (i.e., higher accessibility) at the center of the binding site overlay. This is consistent with higher LYL1 activity in hematopoietic cells. In contrast, both cancer samples show higher relative coverage in the central region, in line with a higher proportion of non-hematopoietic cells contributing to the signal. b) Composite coverage signals around 10,000 GRHL2 binding sites after GC correction. Both cancer samples show a lower central coverage compared to surrounding regions (i.e., higher accessibility), indicating higher activity than in the healthy sample. This is consistent with GRHL2 expression being associated with different cancers.

As pointed out before, exemplary figures of the other report sections, like QC or ichorCNA, can be found in the supplement (Figures S2-S6). The full example report can be found in our GitHub repository.

## Conclusion

Here we propose cfDNA UniFlow, a unified preprocessing pipeline specifically tailored for cfDNA samples. It is an easy-to-use, scalable, and configurable workflow, aiming to set a community standard for enabling accessible and easily sharable future research in the field. In designing our workflow, we aimed at providing a tool that can be used without much computer science background, but with the option to be easily extended by experienced users with their own custom modules , allowing its extension from a standard processing workflow to a full-featured analysis pipeline.

## Data Availability

The data used for generating plots included in this article are available in the European Genome-Phenome Archive at https://ega-archive.org with accession EGAD00001010100.

https://ega-archive.org/datasets/EGAD00001010100

## Availability and Requirements

Project name: **cfDNA-UniFlow**

Project home page: https://github.com/kircherlab/cfDNA-UniFlow

Operating system(s): Linux (64-bit)

Programming language: Python

Other requirements: Mamba or Conda License: MIT

## Additional Files

Supplementary Note: A general overview of the workflow including Supplementary Figures and a quick start guide.

- Supplementary Figure 1: A detailed overview of the workflow components.
- Supplementary Figure 2: Example section of a QC report.
- Supplementary Figure 3: Example report for ichorCNA plot of copy number alterations.
- Supplementary Figure 4: Example of a sample’s global GC bias estimate.
- Supplementary Figure 5: Example report of regional effects of GC bias correction.
- Supplementary Figure 6: Example of a case-control plot.
- Supplementary Figure 7: Comparison of three fragment based GC correction methods.

## Abbreviations

BAM: binary alignment map
cfDNA: cell-free DNA
CNA: copy number alteration
ctDNA: circulating tumor DNA
TF: transcription factor
QC: quality control

## Acknowledgements

We thank current and previous members of the Kircher and Speicher laboratories for helpful discussions and suggestions. Computation has been performed on the HPC for Research cluster of the Berlin Institute of Health at Charité – Universitätsmedizin Berlin.

## Author contributions

Conceptualization: S.R., M.K. and M.R.S.; Data curation: S.R.; Formal analysis: S.R.; Funding acquisition: M.K.; Methodology: S.R.; Project administration: M.K.; Resources: M.K. and M.R.S.; Software: S.R.; Supervision: M.K.; Validation: S.R. and L.B.; Visualization: S.R.; Writing – original draft: S.R.; Writing - review & editing: S.R., L.B., M.R.S. and M.K.

## References

1. Chan AKC, Chiu RWK, Lo YMD, Clinical Sciences Reviews Committee of the Association of Clinical Biochemists. Cell-free nucleic acids in plasma, serum and urine: a new tool in molecular diagnosis. Ann Clin Biochem. 2003; doi: 10.1258/000456303763046030.

2. Lo YM, Zhang J, Leung TN, Lau TK, Chang AM, Hjelm NM. Rapid clearance of fetal DNA from maternal plasma. Am J Hum Genet. 1999; doi: 10.1086/302205.

3. Snyder MW, Kircher M, Hill AJ, Daza RM, Shendure J. Cell-free DNA Comprises an In Vivo Nucleosome Footprint that Informs Its Tissues-Of-Origin. Cell. 2016; doi: 10.1016/j.cell.2015.11.050.

4. Ulz P, Perakis S, Zhou Q, Moser T, Belic J, Lazzeri I, et al. Inference of transcription factor binding from cell-free DNA enables tumor subtype prediction and early detection. Nat Commun. Nature Publishing Group; 2019; doi: 10.1038/s41467-019-12714-4.

5. Ding SC, Lo YMD. Cell-Free DNA Fragmentomics in Liquid Biopsy. Diagn Basel Switz. 2022; doi: 10.3390/diagnostics12040978.

6. Tunc I, Agbor-Enoh S, Valantine H, Thein SL, Pirooznia M. Cfcloud: A Cloud-Based Workflow for Cell-Free DNA Data Analysis. Blood. 2020; doi: 10.1182/blood-2020-138785.

7. Adalsteinsson VA, Ha G, Freeman SS, Choudhury AD, Stover DG, Parsons HA, et al. Scalable whole - exome sequencing of cell-free DNA reveals high concordance with metastatic tumors. Nat Commun. 2017; doi: 10.1038/s41467-017-00965-y.

8. Peneder P, Stütz AM, Surdez D, Krumbholz M, Semper S, Chicard M, et al. Multimodal analysis of cell-free DNA whole-genome sequencing for pediatric cancers with low mutational burden. Nat Commun. 2021; doi: 10.1038/s41467-021-23445-w.

9. Cristiano S, Leal A, Phallen J, Fiksel J, Adleff V, Bruhm DC, et al. Genome-wide cell-free DNA fragmentation in patients with cancer. Nature. 2019; doi: 10.1038/s41586-019-1272-6.

10. Erger F, Nörling D, Borchert D, Leenen E, Habbig S, Wiesener MS, et al. cfNOMe — A single assay for comprehensive epigenetic analyses of cell-free DNA. Genome Med. 2020; doi: 10.1186/s13073-020-00750-5.

11. Shen SY, Singhania R, Fehringer G, Chakravarthy A, Roehrl MHA, Chadwick D, et al. Sensitive tumour detection and classification using plasma cell-free DNA methylomes. Nature. Nature Publishing Group; 2018; doi: 10.1038/s41586-018-0703-0.

12. Chen S, Petricca J, Ye W, Guan J, Zeng Y, Cheng N, et al. The cell-free DNA methylome captures distinctions between localized and metastatic prostate tumors. Nat Commun. Nature Publishing Group; 2022; doi: 10.1038/s41467-022-34012-2.

13. Jung M, Klotzek S, Lewandowski M, Fleischhacker M, Jung K. Changes in concentration of DNA in serum and plasma during storage of blood samples. Clin Chem. 2003; doi: 10.1373/49.6.1028.

14. Lampignano R, Neumann MHD, Weber S, Kloten V, Herdean A, Voss T, et al. Multicenter Evaluation of Circulating Cell-Free DNA Extraction and Downstream Analyses for the Development of Standardized (Pre)analytical Work Flows. Clin Chem. 2020; doi: 10.1373/clinchem.2019.306837.

15. Parpart-Li S, Bartlett B, Popoli M, Adleff V, Tucker L, Steinberg R, et al. The Effect of Preservative and Temperature on the Analysis of Circulating Tumor DNA. Clin Cancer Res Off J Am Assoc Cancer Res. 2017; doi: 10.1158/1078-0432.CCR-16-1691.

16. van Dessel LF, Beije N, Helmijr JCA, Vitale SR, Kraan J, Look MP, et al. Application of circulating tumor DNA in prospective clinical oncology trials – standardization of preanalytical conditions. Mol Oncol. 2017; doi: 10.1002/1878-0261.12037.

17. Abbosh C, Birkbak NJ, Wilson GA, Jamal-Hanjani M, Constantin T, Salari R, et al. Phylogenetic ctDNA analysis depicts early stage lung cancer evolution. Nature. 2017; doi: 10.1038/nature22364.

18. Benjamini Y, Speed TP. Summarizing and correcting the GC content bias in high-throughput sequencing. Nucleic Acids Res. 2012; doi: 10.1093/nar/gks001.

19. Kim CS, Mohan S, Ayub M, Rothwell DG, Dive C, Brady G, et al. In silico error correction improves cfDNA mutation calling. Bioinformatics. 2019; doi: 10.1093/bioinformatics/bty1004.

20. Esfahani MS, Hamilton EG, Mehrmohamadi M, Nabet BY, Alig SK, King DA, et al. Inferring gene expression from cell-free DNA fragmentation profiles. Nat Biotechnol. Nature Publishing Group; 2022; doi: 10.1038/s41587-022-01222-4.

21. Doebley A-L, Ko M, Liao H, Cruikshank AE, Santos K, Kikawa C, et al. A framework for clinical cancer subtyping from nucleosome profiling of cell-free DNA. Nat Commun. 2022; doi: 10.1038/s41467-022-35076-w.

22. Mathios D, Johansen JS, Cristiano S, Medina JE, Phallen J, Larsen KR, et al. Detection and characterization of lung cancer using cell-free DNA fragmentomes. Nat Commun. 2021; doi: 10.1038/s41467-021-24994-w.

23. Markus H, Contente-Cuomo T, Farooq M, Liang WS, Borad MJ, Sivakumar S, et al. Evaluation of pre-analytical factors affecting plasma DNA analysis. Sci Rep. 2018; doi: 10.1038/s41598-018-25810-0.

24. Zheng H, Zhu MS, Liu Y. FinaleDB: a browser and database of cell-free DNA fragmentation patterns. Bioinformatics. Oxford University Press; 2021; doi: 10.1093/bioinformatics/btaa999.

25. Zhang W, Wei L, Huang J, Zhong B, Li J, Xu H, et al. cfDNApipe: a comprehensive quality control and analysis pipeline for cell-free DNA high-throughput sequencing data. Bioinformatics. 2021; doi: 10.1093/bioinformatics/btab413.

26. Mölder F, Jablonski KP, Letcher B, Hall MB, Tomkins-Tinch CH, Sochat V, et al. Sustainable data analysis with Snakemake. F1000Research;

27. Danecek P, Bonfield JK, Liddle J, Marshall J, Ohan V, Pollard MO, et al. Twelve years of SAMtools and BCFtools. GigaScience. 2021; doi: 10.1093/gigascience/giab008.

28. Gaspar JM. NGmerge: merging paired-end reads via novel empirically-derived models of sequencing errors. BMC Bioinformatics. 2018; doi: 10.1186/s12859-018-2579-2.

29. Vasimuddin Md, Misra S, Li H, Aluru S. Efficient Architecture -Aware Acceleration of BWA-MEM for Multicore Systems. 2019 IEEE Int Parallel Distrib Process Symp IPDPS.

30. Andrews S. FASTQC. A quality control tool for high throughput sequence data.

31. Pedersen BS, Quinlan AR. Mosdepth: quick coverage calculation for genomes and exomes. Bioinformatics. 2018; doi: 10.1093/bioinformatics/btx699.

32. Ewels P, Magnusson M, Lundin S, Käller M. MultiQC: summarize analysis results for multiple tools and samples in a single report. Bioinformatics. 2016; doi: 10.1093/bioinformatics/btw354.

33. Zohren F, Souroullas GP, Luo M, Gerdemann U, Imperato MR, Wilson NK, et al. The transcription factor Lyl-1 regulates lymphoid specification and the maintenance of early T lineage progenitors. Nat Immunol. Nature Publishing Group; 2012; doi: 10.1038/ni.2365.

34. Jacobs J, Atkins M, Davie K, Imrichova H, Romanelli L, Christiaens V, et al. The transcription factor Grainy head primes epithelial enhancers for spatiotemporal activation by displacing nucleosomes. Nat Genet. Nature Publishing Group; 2018; doi: 10.1038/s41588-018-0140-x.

35. Chen AF, Liu AJ, Krishnakumar R, Freimer JW, DeVeale B, Blelloch R. GRHL2-Dependent Enhancer Switching Maintains a Pluripotent Stem Cell Transcriptional Subnetwork after Exit from Naive Pluripotency. Cell Stem Cell. 2018; doi: 10.1016/j.stem.2018.06.005.

36. Cocce KJ, Jasper JS, Desautels TK, Everett L, Wardell S, Westerling T, et al. The Lineage Determining Factor GRHL2 Collaborates with FOXA1 to Establish a Targetable Pathway in Endocrine Therapy-Resistant Breast Cancer. Cell Rep. 2019; doi: 10.1016/j.celrep.2019.09.032.

37. Paltoglou S, Das R, Townley SL, Hickey TE, Tarulli GA, Coutinho I, et al. Novel Androgen Receptor Coregulator GRHL2 Exerts Both Oncogenic and Antimetastatic Functions in Prostate Cancer. Cancer Res. 2017; doi: 10.1158/0008-5472.CAN-16-1616.

38. Riethdorf S, Frey S, Santjer S, Stoupiec M, Otto B, Riethdorf L, et al. Diverse expression patterns of the EMT suppressor grainyhead-like 2 (GRHL2) in normal and tumour tissues. Int J Cancer. 2016; doi: 10.1002/ijc.29841.

39. Reese RM, Harrison MM, Alarid ET. Grainyhead-like Protein 2: The Emerging Role in Hormone-Dependent Cancers and Epigenetics. Endocrinology. 2019; doi: 10.1210/en.2019-00213.

40. Kwan EM, Fettke H, Crumbaker M, Docanto MM, To SQ, Bukczynska P, et al. Whole blood GRHL2 expression as a prognostic biomarker in metastatic hormone -sensitive and castration-resistant prostate cancer. Transl Androl Urol. AME Publishing Company; 2021; doi: 10.21037/tau-20-1444.

41. Kumegawa K, Takahashi Y, Saeki S, Yang L, Nakadai T, Osako T, et al. GRHL2 motif is associated with intratumor heterogeneity of cis-regulatory elements in luminal breast cancer. Npj Breast Cancer. Nature Publishing Group; 2022; doi: 10.1038/s41523-022-00438-6.

