## Supplement for "cfDNA UniFlow: A unified preprocessing pipeline for cell-free DNA data from liquid biopsies"

cfDNA UniFlow is a unified, standardized, and ready-to-use workflow for processing whole genome sequencing (WGS) cfDNA samples from liquid biopsies. It includes essential steps for pre-processing raw cfDNA samples, quality control and reporting. Additionally, several optional utility functions like GC bias correction and estimation of copy number state are included. Finally, we provide specialized methods for extracting coverage derived signals and visualizations comparing cases and controls. Figure S1 gives a detailed overview of the workflow.

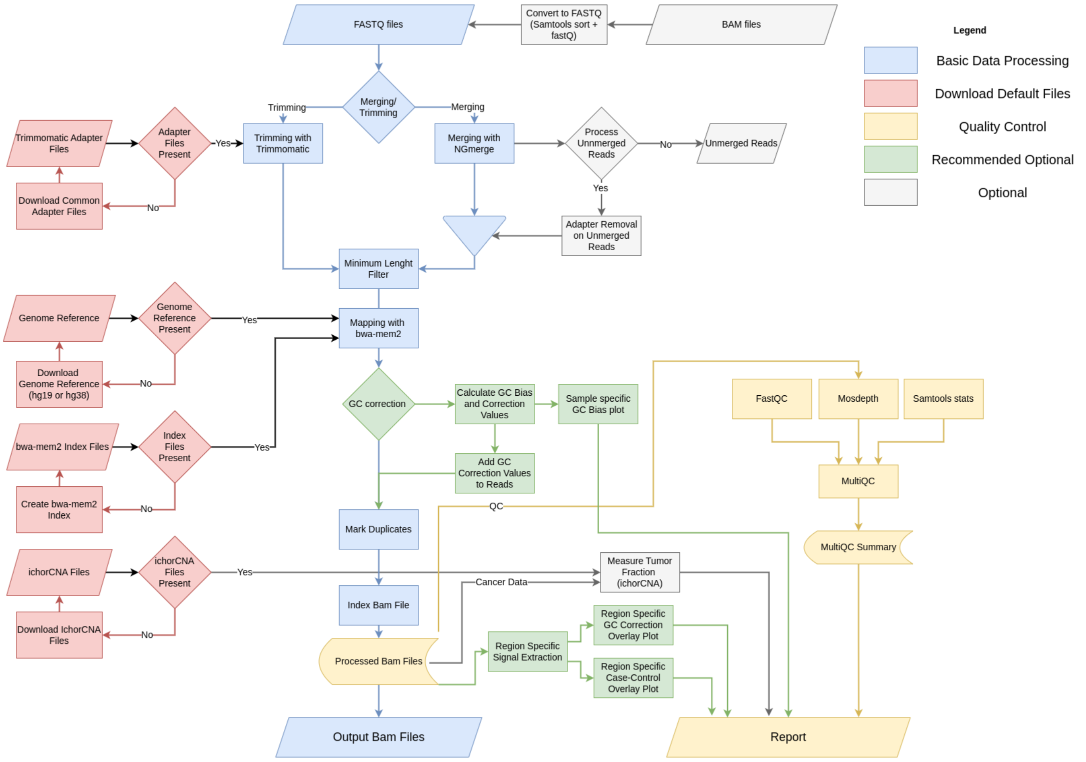


**Figure S1: Overview of cfDNA Uniflow.** Functionalities are color coded by task. Red boxes represent rules for the automatic download of public resources. Grey boxes are optional steps. Blue boxes contain the core functionalty of cfDNA Uniflow. Green boxes are optional, but highly recommended steps and yellow boxes summarize the Quality Control and reporting steps.

#### Authors


- Sebastian Röner (@sroener)

### Table of Contents


- cfDNA UniFlow: A unified preprocessing pipeline for cell-free DNA from liquid biopsies
  - 1 Dependencies
  - 2 Functional summary
    - 2.1 Raw data processing
    - 2.2 Quality control
    - 2.3 Signal extraction and sequence analysis functionality
    - 2.4 Report
    - 2.5 Comparison of fragment based GC bias correction methods for cfDNA
    - 2.6 Notes on resource requirements
  - 3 Quick-start Guide
    - 3.1 Setup
      - Step 1: Obtain a copy of this workflow
      - Step 2: Install Snakemake
      - Step 3: Activate conda environment
    - 3.2 Integration test
      - Step 1: Download test files
      - Step 2: Check config files
      - Step 3: Executing the workflow
    - 3.3 Running the workflow with the data used in the manuscript
      - Step 1: Download files from EGA
      - Step 2: Check manuscript config files
      - Step 3: Executing the workflow
      - Step 4: Reproducing figures 2 and 3
    - 3.4 Running the workflow with your own data
      - Step 1: Configure Workflow
      - Step 2: Execute workflow
      - Step 3: Investigate results
  - 4 Contribution guidelines
  - 5. References

### 1 Dependencies

To minimize conflicts between packages, all dependencies for the workflow rules are managed via separate conda environments located in `./workflow/envs`. They get automatically installed and used when Snakemake is executed with the `--use-conda` flag. This is the recommended way of executing the workflow.

The only exception is NGmerge, a read merging and adapter removal program, which is included in the GitHub repository due to an outdated bioconda recipe. The NGmerge executable was downloaded and compiled as described in the official documentation of NGmerge v0.3 and is located in the scripts directory (./workflow/scripts/NGmerge). Additionally, we provide an adjusted quality profile for Phred+33 scores of Illumina 1.8+, which ranges from 0 to 41 instead of 0 to 40 in earlier versions. The quality profile file was modified by duplicating the last column and appending it as a new column.

### 2 Functional summary

#### 2.1 Raw data processing

The core functionality of cfDNA UniFlow is the processing of Whole Genome Sequencing (WGS) data from liquid biopsies. Input data is expected in either FASTQ or BAM format. If a BAM file was provided, it gets converted to FASTQ files using SAMtools. Afterwards, several steps for improving read quality and preparation for mapping are executed, for which two options are provided. Either the recommended merging of reads with NGmerge, which removes adapters and corrects sequencing errors, or trimming of adapters with Trimmomatic. In both cases, results are filtered for a specified minimum read length. Remaining reads are mapped to a reference genome via bwa-mem2. If NGmerge was used for adapter removal, it is possible to include reads in mapping for which only adapters were removed when merging was not possible due to no sufficient overhang between read pairs. The core processing is finalized by marking duplicates and creating a bam index with SAMtools markdup and index. Processed reads are then submitted for Quality Control and optional steps.

#### 2.2 Quality control

In the quality control step, general post-alignment statistics and graphs are calculated for each sample via SAMtools stats and FastQC. Additionally, sample-wide coverage statistics and coverage at different genomic regions are calculated with Mosdepth, a fast BAM/CRAM depth calculation tool for WGS, exome, or targeted sequencing. The QC results are aggregated in HTML report via MultiQC.

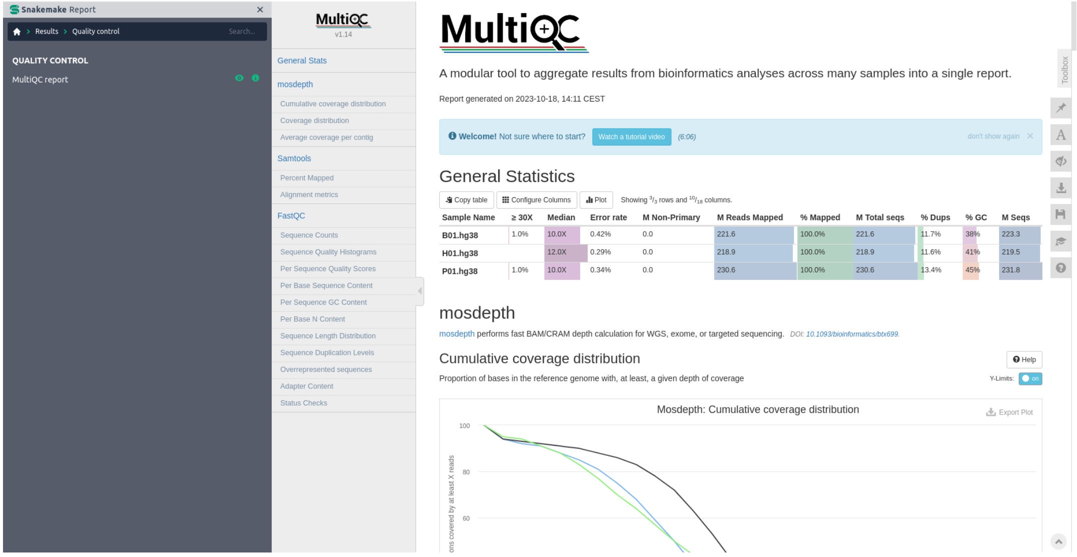


**Figure S2: Example section of a QC report.** The QC report contains post-alignment statistics from SAMtools stats, FastQC and Mosdepth.

#### 2.3 Signal extraction and sequence analysis functionality

In addition to the preprocessing and quality control functionality, cfDNA UniFlow contains some sequence analysis functions. The first is the widely used tool ichorCNA, which can be used for predicting copy number alteration (CNA) states across the genome. Further, it uses this information for estimating tumor fractions in cfDNA samples. By default, we use the recommended settings for ichorCNA, including profiles provided in the ichorCNA repository. However, it is possible to specify custom profiles and parameters in the configuration file.

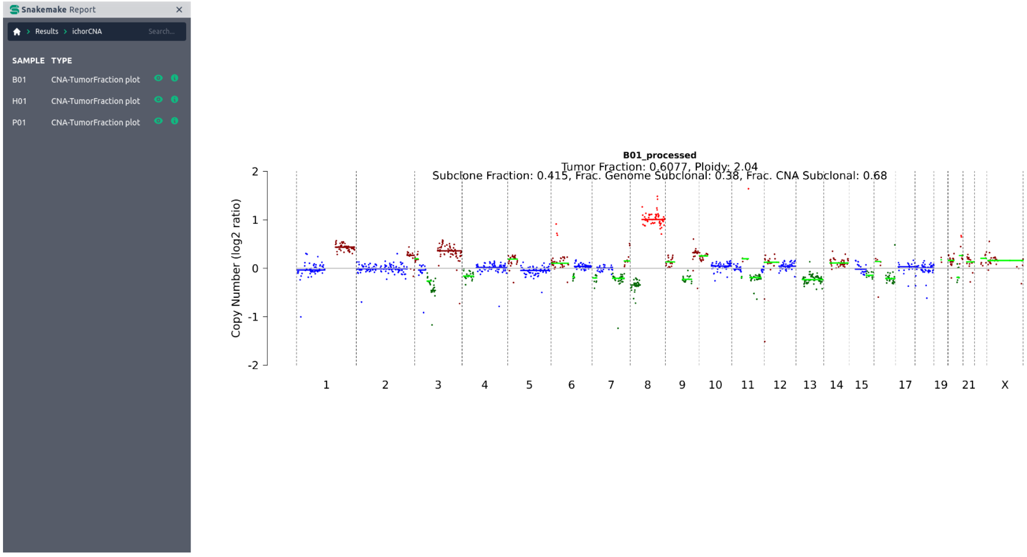


**Figure S3: ichorCNA plot of copy number alterations.** The plot shows genome wide annotation of estimated copy number alterations, tumor fraction and other parameters used by ichorCNA. This example was generated using a breast cancer sample with an average GC content of 38%.

The second utility function is our inhouse GC bias estimation method. It can not only be used for estimating fragment length and GC-content dependent technical biases, but also includes the option of attaching correction values to the reads. These can be used downstream for a wide variety of signal extraction methods, while preserving the original read coverage patterns. Additionally, it is possible to include a visualization of the estimated biases in the report.

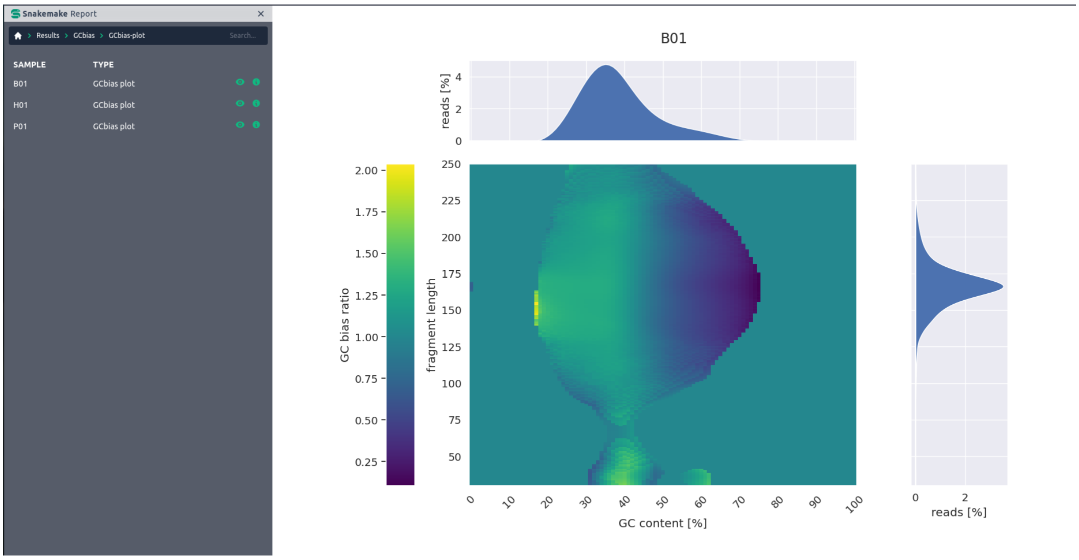


**Figure S4: Global GC bias estimate.** The global GC bias was estimated using ratio between observed and expected fragments stratified by GC content [%] and fragment length [bp].This example was generated using a breast cancer sample with an average GC content of 38%.

Building on the GC bias estimation, we provide a method for extracting coverage derived signals around user defined regions. The resulting signals can be visualized for comparing biased vs corrected states and for comparing cases against controls.

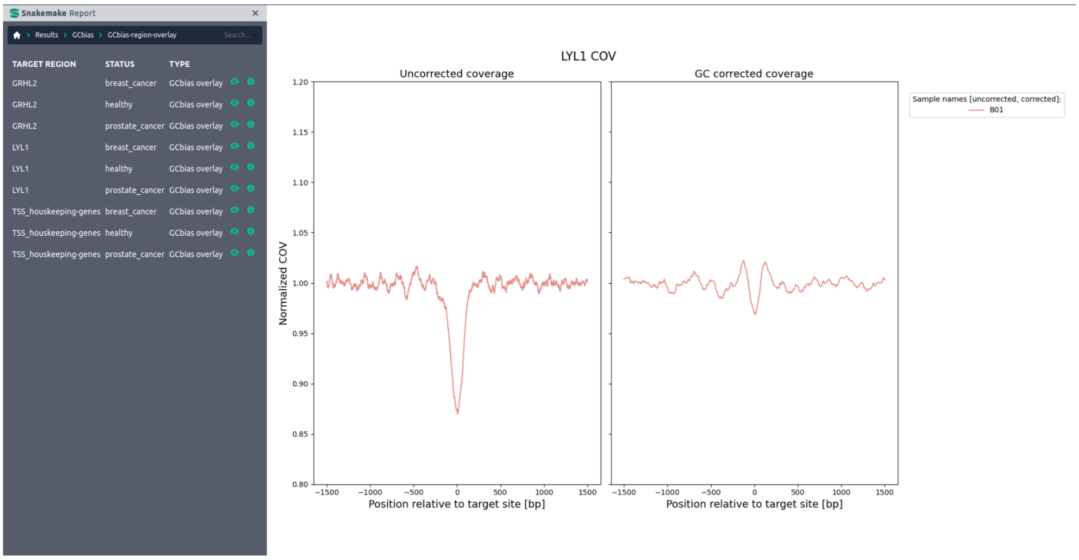


**Figure S5: Regional effects of GC bias correction.** Composite coverage signal of 10000 LYL1 binding sites before and after GC bias correction. Accessibility of the binding sites, indicated by lower coveragem would have been overestimated before correction. This example was generated using a breast cancer sample with an average GC content of 38%.

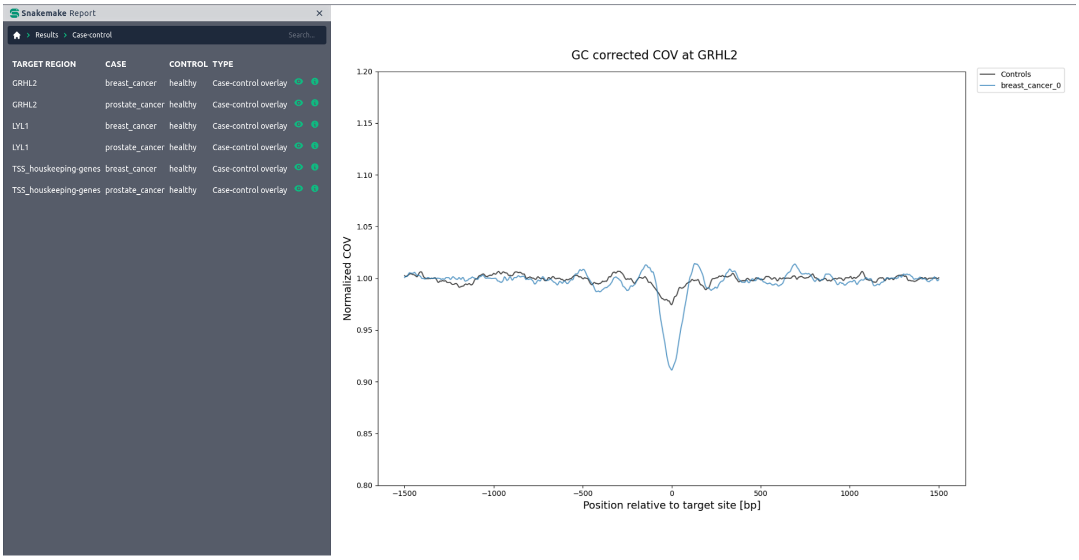


**Figure S6: Case-control plot.** GC corrected composite coverage signal of 10000 GRHL2 binding sites in a healthy control and a breast cancer sample. Lower central coverage in the cancer sample is consistent with increased GRHL2 activity in many cancer types.

#### 2.4 Report

Finally, all results and summary statistics for the specified samples are aggregated in one report, making a wide variety of information easily accessible. The report file is generated using Snakemake's report feature. After the workflow finished, the report can be generated by executing Snakemake –configfile <CONFIGFILE> --snakefile <SNAKEFILE> --report <REPORTNAME>.html. For better report performance, it is recommended to use --report <REPORTNAME>.zip, which creates a zipped directory structure with the needed information instead of saving it in the HTML file itself. More information on reporting can be found in the official Snakemake documentation.

**Note:** An example report can be found here

#### 2.5 Comparison of fragment based GC bias correction methods for cfDNA

The figure below shows a comparison of fragment based GC bias correction methods for cfDNA. We compared our inhouse method (ID: cfDNA Uniflow) with the methods from GCparagon [1] and scores extracted from the Griffin workflow [2]. Expected and measured fragment distributions were sampled from 1e5 random regions of 1 kilobase (kb) width, covering 1e8 base pair (bp) in total. The expected fragment distribution is based on all possible fragments of size 20 to 550 bp and their respective GC content based on the human genome reference hg38. The measured fragment distribution is based on the actual fragments in the cfDNA sample.

**A)**

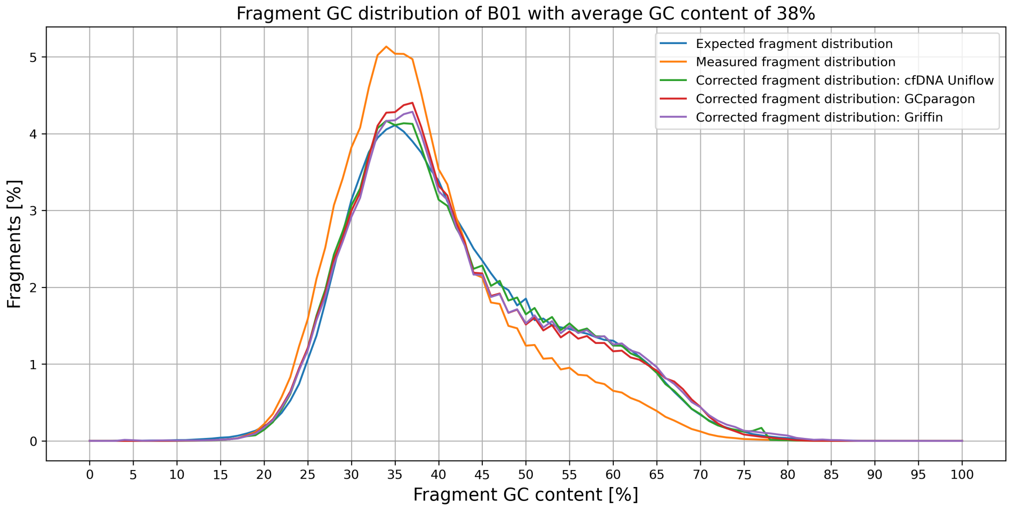

Fragment GC distribution of B01 with average GC content of 38%
Expected fragment distribution
Measured fragment distribution
Corrected fragment distribution: cfDNA Uniflow
Corrected fragment distribution: GCparagon
Corrected fragment distribution: Griffin
Fragments [%]
Fragment GC content [%]


**B)**

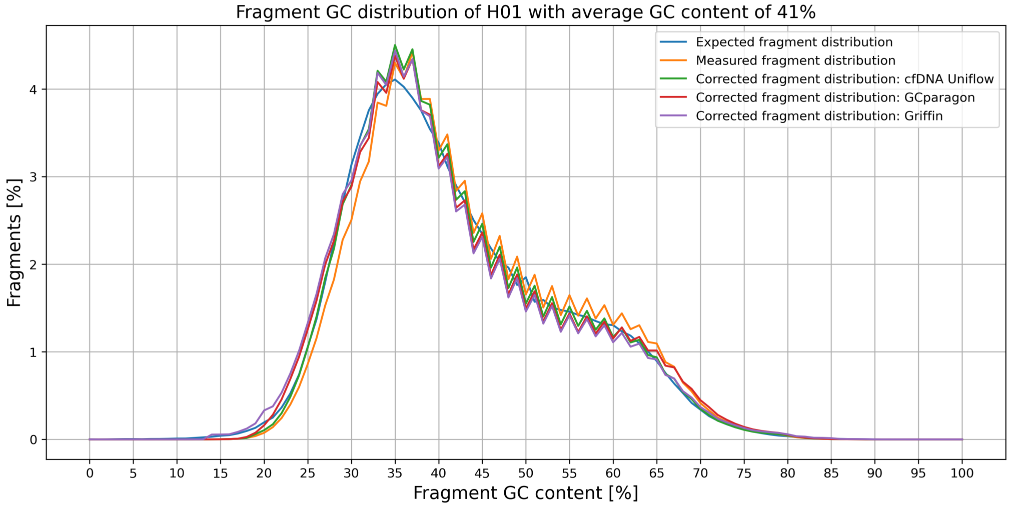

Fragment GC distribution of H01 with average GC content of 41%
Expected fragment distribution
Measured fragment distribution
Corrected fragment distribution: cfDNA Uniflow
Corrected fragment distribution: GCparagon
Corrected fragment distribution: Griffin
Fragments [%]
Fragment GC content [%]


**C)**

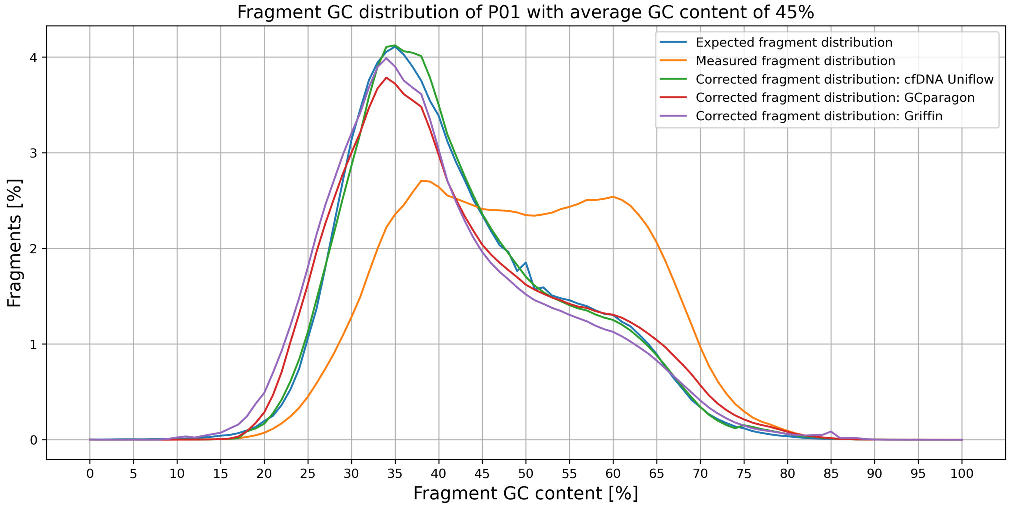

Fragment GC distribution of P01 with average GC content of 45%
Expected fragment distribution
Measured fragment distribution
Corrected fragment distribution: cfDNA Uniflow
Corrected fragment distribution: GCparagon
Corrected fragment distribution: Griffin
Fragments [%]
Fragment GC content [%]

**Figure S7: Comparison of three fragment-based GC bias correction methods for cfDNA.** The expected fragment distribution is based on all possible fragments of size 20 to 550 bp and their respective GC content based on the human genome reference hg38. The measured fragment distribution is based on the actual fragments in the cfDNA sample. Both distributions were sampled from 1e5 random regions of 1 kilobase (kb) width, covering 1e8 base pair (bp) in total. Corrected fragment distributions were calculated by multiplying the fragment counts with GC correction factors of the respective methods. Subfigure A) show the effects for the tested sample B01 (38% average GC), B) for sample H01 (41% average GC) and C) for sample P01 (47% average GC).

#### 2.6 Notes on resource requirements

- The index creation of bwa-mem2 is resource intensive:

```
# Indexing the reference sequence (Requires 28N GB memory where N is the size of the reference sequence).
./bwa-mem2 index [-p prefix] <in.fasta>
Where
<in.fasta> is the path to reference sequence FASTA file and
<prefix> is the prefix of the names of the files that store the resultant index. Default is in.FASTA.
```

More information can be found in the documentation or this issue.

- bwa-mem2 mem uses around 4GB memory per thread.

### 3 Quick-start Guide

#### 3.1 Setup

The goal of cfDNA Uniflow is to provide essential and standardized preprocessing steps in a reproducible and scalable way, with the option to include additional steps as needed. Therefore, we encourage users to use this workflow as a template and build their own analysis on top of it. The following steps guide you through the setup of the workflow.

**Note 1:** If you use this workflow in a paper, don't forget to give credits to the authors by citing the URL of this (original) repository. **Note 2:** If you want to contribute to the development of cfDNA UniFlow, please follow the guidelines in section 4.

##### Step 1: Obtain a copy of this workflow

1. Create a new GitHub repository using this workflow as a template.
2. Clone the newly created repository to your local system, into the place where you want to perform the data analysis.

###### Step 2: Install Snakemake

After successful installation, set up an environment for Snakemake. This can be done by executing the following command:

```
conda create -c bioconda -c conda-forge -n snakemake snakemake
```

The environment can be activated via the `conda activate snakemake` command.

For installation details, see the instructions in the Snakemake documentation.

###### Step 3: Activate conda environment

Activate the conda environment:

```
conda activate snakemake
```

##### 3.2 Integration test

Our goal in developing cfDNA UniFlow is to provide a scalable, configurable, and easy-to-use workflow specifically tailored towards the processing of cfDNA samples. Users only need to provide sequencing information in FASTQ or BAM format and optionally modify the configuration file to their needs. Here we provide an example with a small input file for testing the workflows functionality.

**Note:** As this is an integration test, the generated data is not intended to generate meaningful results. This test is only meant to check if the workflow is running without errors.

###### Step 1: Download test files

First, you should create the expected directory for the test files:

```
mkdir -p resources/testsample/
```

Afterwards, download the files located on this webserver:

```
curl -L -o resources/testsample/testsample_hg19_1x_chr20-22.bam
https://kircherlab.bihealth.org/download/cfDNA-
testSample/testsample_hg19_1x_chr20-22.bam
```

###### Step 2: Check config files

There are three files that are used in this example:

- config/test-config.yaml

- config/test-samples.tsv
- config/test-regions.tsv

**Note**: These files are configured for a quick integration test and shouldn't be edited.

###### Step 3: Executing the workflow

The last step is executing the Workflow. For this you need to be in the root directory of the cloned repository.

Test the configuration with a dry-run:

```
snakemake --use-conda --configfile config/test-config.yaml -n
```

The workflow is executed locally with $N cores via:

```
snakemake --use-conda --configfile config/test-config.yaml --cores $N
```

For cluster execution, read the guidelines in the Snakemake documentation.

##### 3.3 Running the workflow with the data used in the manuscript

###### Step 1: Download files from EGA

The data used in the manuscript is available on the European Genome-Phenome Archive (EGA) under the accession number EGAD00001010100. For the download, you need to have an EGA account and request access via the EGA website. Afterwards, the data can be downloaded using the EGA download client.

After downloading the data, the files need to be placed in the directory `resources/manuscript_samples/` relative to the repository root. If the directory does not exist, create it with the following command:

```
mkdir -p resources/manuscript_samples/
```

###### Step 2: Check manuscript config files

We provide a configuration file for the manuscript data in the `supplement/manuscrip_analysis` directory. The directory contains the following files:

- manuscript_config.yaml
- manuscript_samples.tsv
- cfDNA_Uniflow_figure2-3.ipynb

**Note:** Please check the paths/file names in the `manuscript_samples.tsv` file and adjust them if necessary.

###### Step 3: Executing the workflow

This step is similar to the integration test, but with the manuscript data.

Test the configuration with a dry-run:

```
snakemake --use-conda --configfile supplement/manuscrip_analysis/manuscript_config.yaml -n
```

The workflow is executed locally with $N cores via:

```
snakemake --use-conda --configfile supplement/manuscrip_analysis/manuscript_config.yaml --cores $N
```

If possible, we highly encourage the execution of the workflow in a cluster environment. The detailed configuration depends on the available infrastructure. More information can be found in the respective subsection of the Snakemake documentation.

###### Step 4: Reproducing figures 2 and 3

To reproduce the figures 2 and 3 from the manuscript, you need to execute the Jupyter notebook `cfDNA_Uniflow_figure2-3.ipynb` located in the `supplement/manuscript_analysis` directory. The notebook contains the code for generating the figures and is based on the results generated by the workflow.

#### 3.4 Running the workflow with your own data

After running the integration test, you can start using the workflow with your own data. For this, you need to modify the configuration files and provide your own data.

###### Step 1: Configure Workflow

Configure the workflow according to your needs via editing the files in the `config/` folder. Adjust `config.yaml` according to the included comments to configure the workflow execution. The most important options are the path to the respective `samples.tsv` and `regions.tsv` files. All other default files are either included, or will be downloaded and generated automatically.

The `samples.tsv` file should contain the following columns:

```
# Identifier of the cohort/experiment
ID: string
# Sample name/identifier
sample: string
# Path to the BAM file (for reprocessing); insert "-" if FASTQ files
are used
bam: string
# Path to the R1 FASTQ file; insert "-" if bam is used
fq1: string
# Path to the R2 FASTQ file; insert "-" if bam is used
fq2: string
# Name of the target genome build (hg19 or hg38)
genome_build: string
# Name of the library prep (e.g. ThruPLEX DNA-seq)
library_name: string
# Platform used for sequencing (e.g. Illumina NextSeq 500)
platform: string
# Status of the sample (e.g. Case/Control or Healthy/Cancer)
status: string
# Additional information that should be included in read group ID
(e.g. condition)
info: string
```

If FASTQ files are used, either single-end or paired-end sequencing can be specified, depending on how many FASTQ files are provided. If BAM files are used, the FASTQ columns should be filled with "-" and vice versa.

The `regions.tsv` file should contain the following columns:

```
# region name/identifier
target: string
# path to the bed files for regions of interest
path: string
```

**Note:** All three files will be automatically validated before workflow execution.

**Step 2: Execute workflow**

Activate the conda environment:

```
conda activate snakemake
```

Test your configuration by performing a dry-run with `<CONFIGFILE>` representing the path to your modified config file via:

```
snakemake --use-conda --configfile <CONFIGFILE> -n
```

Execute the workflow locally using `$N` cores:

```
snakemake --use-conda --configfile <CONFIGFILE> --cores $N
```

See the Snakemake documentation on workflow execution and execution in cluster environments for further details.

###### Step 3: Investigate results

After successful execution, you can create a self-contained interactive HTML report with all results via:

```
snakemake --configfile <CONFIGFILE> --report report.html
```

This report can, e.g., be forwarded to your collaborators.

A functional description of reporting can be found in section 2.4.

An example report in zip format can be found in the supplement directory. For viewing, the zip file needs to be extracted.

#### 4 Contribution guidelines

In case you want to contribute to the development of cfDNA UniFlow and its core functionalities, please follow these steps:

1. Fork the original repo to a personal or lab account.
2. Clone the fork to your local system, to a different place than where you ran your analysis.
3. Modify the code according to the intended changes.
4. Commit and push your changes to your fork.
5. Create a pull request against the original repository.

#### 5. References

[1] Spiegl B, Kapidzic F, Röner S, Kircher M, Speicher MR. GCparagon: evaluating and correcting GC biases in cell-free DNA at the fragment level. NAR Genomics Bioinforma. 2023; doi: 10.1093/nargab/lqad102.

[2] Doebley A-L, Ko M, Liao H, Cruikshank AE, Santos K, Kikawa C, et al.. A framework for clinical cancer subtyping from nucleosome profiling of cell-free DNA. Nat Commun. 2022; doi: 10.1038/s41467-022-35076-w.
